# Real-World Evaluation of Large Language Models in Healthcare (RWE-LLM): A New Realm of AI Safety & Validation

**DOI:** 10.1101/2025.03.17.25324157

**Authors:** Meenesh Bhimani, Alex Miller, Jonathan D. Agnew, Markel Sanz Ausin, Mariska Raglow-Defranco, Harpreet Mangat, Michelle Voisard, Maggie Taylor, Sebastian Bierman-Lytle, Vishal Parikh, Juliana Ghukasyan, Rae Lasko, Saad Godil, Ashish Atreja, Subhabrata Mukherjee

## Abstract

**Background:** The deployment of artificial intelligence (AI) in healthcare necessitates robust safety validation frameworks, particularly for systems directly interacting with patients. While theoretical frameworks exist, there remains a critical gap between abstract principles and practical implementation. Traditional LLM benchmarking approaches provide very limited output coverage and are insufficient for healthcare applications requiring high safety standards.

**Objective:** To develop and evaluate a comprehensive framework for healthcare AI safety validation through large-scale clinician engagement.

**Methods:** We implemented the RWE-LLM (Real-World Evaluation of Large Language Models in Healthcare) framework, drawing inspiration from red teaming methodologies while expanding their scope to achieve comprehensive safety validation. Our approach emphasizes output testing rather than relying solely on input data quality across four stages: pre-implementation, tiered review, resolution, and continuous monitoring. We engaged 6,234 US licensed clinicians (5,969 nurses and 265 physicians) with an average of 11.5 years of clinical experience. The framework employed a three-tier review process for error detection and resolution, evaluating a non-diagnostic AI Care Agent focused on patient education, follow-ups, and administrative support across four iterations (pre-Polaris and Polaris 1.0, 2.0, and 3.0).

**Results:** Over 307,000 unique calls were evaluated using the RWE-LLM framework. Each interaction was subject to potential error flagging across multiple severity categories, from minor clinical inaccuracies to significant safety concerns. The multi-tiered review system successfully processed all flagged interactions, with internal nursing reviews providing initial expert evaluation followed by physician adjudication when necessary. The framework demonstrated effective throughput in addressing identified safety concerns while maintaining consistent processing times and documentation standards. Systematic improvements in safety protocols were achieved through a continuous feedback loop between error identification and system enhancement. Performance metrics demonstrated substantial safety improvements between iterations, with correct medical advice rates improving from ∼80.0% (pre-Polaris), to 96.79% (Polaris 1.0), to 98.75% (Polaris 2.0) and 99.38% (Polaris 3.0). Incorrect advice resulting in potential minor harm decreased from 1.32% to 0.13% and 0.07%, and severe harm concerns were eliminated (0.06% to 0.10% and 0.00%).

**Conclusions:** The successful nationwide implementation of the RWE-LLM framework establishes a practical model for ensuring AI safety in healthcare settings. Our methodology demonstrates that comprehensive output testing provides significantly stronger safety assurance than traditional input validation approaches used by horizontal LLMs. While resource-intensive, this approach proves that rigorous safety validation for healthcare AI systems is both necessary and achievable, setting a benchmark for future deployments.

## 1. Introduction

The rapid advancement of artificial intelligence (AI) in healthcare presents unprecedented opportunities for improving patient outcomes and addressing critical challenges such as the ongoing clinician shortage. However, it also introduces novel safety considerations that must be rigorously addressed before widespread deployment. While existing frameworks provide theoretical foundations for AI safety validation, a significant gap remains between abstract principles and practical implementation, particularly for AI systems designed to interact directly with patients. Current approaches often lack the scale, diversity, and structured validation processes necessary to ensure reliable safety outcomes across varied clinical contexts.

Current approaches to LLM safety in healthcare focus predominantly on input testing and limited benchmark evaluations, which cover only a small fraction of possible model outputs. These methods provide insufficient safety assurance for AI systems designed for direct patient interaction. The RWE-LLM framework addresses this critical gap by implementing comprehensive output testing across diverse clinical scenarios, expanding upon red teaming methodologies to achieve unprecedented coverage of potential AI responses.

Conventional LLM safety approaches—including training data curation, prompt engineering, and post-processing filters—provide insufficient safety guarantees for healthcare applications. These methods fail to capture the full complexity of clinical communication and cannot anticipate the diverse ways patients might interact with AI systems.

Unlike horizontal LLMs designed for general-purpose applications, healthcare AI systems require specialized safety validation that addresses their unique deployment contexts. Horizontal LLMs are typically evaluated using general benchmarks that sample only a tiny fraction of possible outputs, providing limited safety assurance. By contrast, the approach described in this paper implements comprehensive output testing rather than relying on these limited benchmarks. No horizontal LLM currently employs such extensive output validation, making this approach distinctive in its focus on comprehensive safety coverage across diverse clinical scenarios.

This paper presents a comprehensive safety validation framework, RWE-LLM (Real-World Evaluation of Large Language Models in Healthcare), implemented through the engagement of over 6,200 clinicians. The framework integrates robust error detection and resolution protocols while demonstrating scalable approaches to safety assurance. By combining clinician evaluation, error management systems, and quality control processes, the RWE-LLM framework establishes a benchmark for AI safety validation. Addressing both safety and scalability, this framework provides a critical step toward bridging the gap between the potential of AI and real-world healthcare transformation.

## 2. Background

The development of safety frameworks for healthcare AI systems has evolved significantly in recent years, moving from theoretical and technical considerations to more comprehensive approaches that encompass ethical, social, and operational dimensions.

### 2.1 Evolution of Healthcare AI Safety Frameworks

Early approaches to healthcare AI safety focused primarily on technical validation and performance metrics. However, recent frameworks have expanded to include broader considerations of safety and responsibility. Davahli et al. proposed the Safety Controlling System (SCS), a three-level multi-attribute value model that evaluates AI safety through six key attributes, 13 second-level attributes, and 78 third-level measurable attributes.^1^ This hierarchical approach demonstrates the complexity of safety considerations in healthcare AI, encompassing elements from safety policies and clinician incentives to training requirements and control mechanisms.

The growing recognition of stakeholder perspectives has led to more inclusive frameworks. Sujan et al. conducted a comprehensive stakeholder analysis, revealing diverse perceptions of AI safety and assurance across patients, hospital staff, technology developers, and regulators.^2^ Their findings emphasize the need for a systems-level approach that extends beyond technical considerations to address broader socio-technical implications, including the critical patient-clinician relationship.

A particularly challenging aspect of healthcare AI safety is the “black box” problem, where complex algorithms like deep neural networks make predictions without providing transparent explanations for their decisions.^3^ This opacity complicates validation efforts and raises concerns about accountability, particularly in high-stakes clinical contexts where understanding the reasoning behind recommendations is essential for both providers and patients.

Morley et al. further expanded this perspective by proposing a framework that evaluates not only safety but also acceptability and efficacy of AI systems in healthcare.^4^ Their work emphasizes the interconnected nature of these aspects and the need for holistic evaluation approaches. This evolution toward comprehensive frameworks is also evident in the work of Ellahham et al., who explored both opportunities and challenges in healthcare safety contexts.^5^

### 2.2 Implementation Science and Practical Considerations

Recent frameworks have increasingly focused on practical implementation methodologies. Reddy et al. developed an evaluation framework specifically designed to guide the implementation of AI systems in healthcare settings, emphasizing the importance of structured approaches to system deployment.^6^ This work was complemented by Higgins and Madai, who proposed a practical framework for AI product development in healthcare, bridging the gap between theoretical safety principles and real-world application.^7^

The importance of trust in healthcare AI systems has been highlighted by de-Manuel-Vicente et al., who developed a framework for trustworthy AI with practical code pipelines.^8^ This technical perspective is balanced by Macrae’s work on managing risk and resilience in autonomous and intelligent systems, which emphasizes the dynamic nature of safety in healthcare AI applications.^9^

Recent systematic reviews have identified several established frameworks that guide AI implementation in healthcare settings. The Consolidated Framework for Implementation Research (CFIR) has been successfully adapted to analyze barriers and facilitators specific to AI implementation, while the Nonadoption, Abandonment, Scale-up, Spread, and Sustainability (NASSS) framework has proven valuable for evaluating the complexity of AI technologies and their integration into healthcare systems.^10^ Additional frameworks like SALIENT (Staged Application of Learning and Implementation of New Evidence and Technology) have been developed specifically for healthcare technology implementation, with explicit consideration of AI deployment challenges.^11^ Key implementation considerations identified across these frameworks include the importance of leadership support and organizational culture, stakeholder engagement throughout the implementation process, resource allocation, workflow integration, and the need for continuous monitoring and improvement.^12–17^.

### 2.3 LLM Benchmarking Limitations and Red Teaming Approaches

LLM evaluations in healthcare have been criticized for an overemphasis on accuracy. Bedi et al. report that nearly 95.4% of assessments focus on accuracy while only 15.8% address fairness, bias, or toxicity; merely 5% incorporate real patient care data.^18^ Hager et al. and Liu et al. document that simulations using curated clinical datasets expose deficits in diagnostic accuracy and guideline adherence, highlighting shortcomings in current output coverage.^19,20^

These benchmarking limitations are particularly problematic for healthcare applications where safety and reliability are paramount. Traditional benchmarks provide very low coverage of possible outputs, testing only a minute fraction of the potential response space that an LLM might generate when deployed in real-world clinical settings. This limited coverage creates significant blind spots in safety validation, especially for systems designed to interact directly with patients across diverse clinical scenarios.

Current approaches to LLM safety in healthcare largely focus on three main strategies, each with significant limitations. First, training data curation attempts to ensure safety by carefully selecting high-quality, medically accurate inputs. However, this approach cannot guarantee that the model will generate safe outputs in all scenarios, as LLMs can combine information in unpredictable ways. Second, prompt engineering techniques like constitutional AI and instruction fine-tuning aim to align model behavior with safety guidelines, but these methods often struggle with complex medical nuances and context-dependent safety considerations. Third, post-processing filters screen outputs for harmful content but typically rely on keyword matching or simplistic classifiers that miss subtle clinical safety issues. These approaches fundamentally represent “input testing” paradigms that fail to comprehensively validate the actual outputs that patients might receive in diverse clinical contexts.

A common request from healthcare stakeholders when evaluating AI systems is “tell me what your AI is trained on so I know if it is safe.” This reflects a widespread but incomplete understanding of LLM safety, assuming that safety is primarily determined by training data quality. While high-quality medical training data is necessary, it is insufficient to guarantee safety. The complexity of generative AI systems means they can produce novel combinations of information not present in training data, including potentially harmful outputs even when trained exclusively on vetted medical content. Safety cannot be inferred solely from training inputs but must be validated through comprehensive output testing across diverse clinical scenarios. The RWE-LLM framework addresses this fundamental limitation by directly evaluating what matters most: the actual responses patients would receive in real-world interactions.

Red teaming methods have been adapted to broaden testing in medical settings. Chang et al. demonstrate that interdisciplinary expert evaluations, which target safety, privacy, hallucinations, and bias, can uncover hidden vulnerabilities.^21^ Ness et al. introduce the MedFuzz method to probe model robustness to input variations, and Pfohl et al. propose a multifactorial framework to assess health equity biases.^22,23^ In doing so, these studies expand the testing scope from mere question-answering to include complex, real-world clinical applications. These red teaming approaches provided crucial inspiration for the RWE-LLM framework, particularly their adversarial testing methodology and focus on comprehensive safety evaluation rather than narrow performance metrics.

While these red teaming approaches represent important advances in healthcare AI evaluation, they typically remain limited in scale and clinical diversity. The RWE-LLM framework builds upon these foundations by dramatically expanding testing coverage through large-scale clinician engagement across diverse specialties and practice settings. This approach fundamentally shifts the validation paradigm from input testing to comprehensive output testing, acknowledging that even well-trained LLMs may produce unsafe responses in real-world healthcare scenarios.

Finally, a fundamental difference between our approach and those used by horizontal LLMs lies in testing methodology. Horizontal LLMs are typically evaluated on standardized benchmarks that collectively represent an extremely small sample of potential outputs. For example, a typical medical subset of MMLU might contain only a few hundred questions.^24^ Our framework evaluated over 307,000 unique clinical interactions. This represents orders of magnitude greater coverage, particularly for specialized healthcare contexts. Additionally, horizontal LLMs focus primarily on accuracy rather than safety, with limited attention to the potential harms of incorrect outputs in high-stakes settings. The RWE-LLM framework explicitly addresses this gap by focusing on comprehensive output safety testing, providing a level of validation not currently implemented by any horizontal LLM provider.

### 2.4 Governance and Regulatory Perspectives

Governance frameworks have emerged as a critical component of healthcare AI safety. Reddy et al. proposed a comprehensive governance model for healthcare AI applications, while Larson et al. focused specifically on regulatory frameworks for AI-based diagnostic imaging algorithms.^25,26^ These governance approaches are particularly important given the potential for bias and identified clinical safety issues.^27^

The role of regulation in ensuring AI safety has been further explored by Kiseleva, who questioned whether performance transparency and accountability measures are sufficient for AI medical devices.^28^ This regulatory perspective is complemented by work from Ross and Spates on quality considerations in healthcare AI, namely the need to overcome the “black box” problem of AI through greater transparency and understanding.^29^

### 2.4 Current Challenges and Future Directions

Despite significant progress, several important gaps remain in current frameworks. Cresswell et al. highlight the importance of not “reinventing the wheel” when evaluating AI in clinical settings, suggesting that existing evaluation methodologies can be adapted for AI systems.^30^ Crossnohere et al. conducted a comprehensive content analysis of AI guidelines in medicine, revealing inconsistencies and gaps in current approaches, foremost among these being the lack of engagement in oversight of key stakeholders.^31^ Despite having at least a dozen well-developed frameworks, the literature provides little or inconsistent real-world evidence in relation to their impact and application.^30^

Integration challenges remain a significant concern, as identified in analyses of emerging safety issues.^32^ Cultural adaptation represents another critical gap, with researchers emphasizing the need for responsible AI frameworks that can adapt to different healthcare contexts.^33^

The resource requirements for implementing comprehensive safety frameworks, particularly in resource-constrained settings, remain poorly addressed in current literature. Bates et al. identified this as a key challenge in their scoping review of AI’s potential to improve patient safety.^34^ Additionally, long-term validation and monitoring of AI systems in healthcare settings requires further attention.^35^ Hence, Longhurst et al. advocated the central need for including implementation science principles and measurement for clinical efficacy at the point of care where most of the variability lies.^36^ However, there is no overarching framework that can guide simulated clinical validation before deployment at the point of care.

## 3. Methods

The Hippocratic AI Care Agent represents a novel AI system designed to act as a scalable digital healthcare assistant focused on non-diagnostic patient support. This AI system was developed to interact directly with patients across various healthcare contexts, including patient education, post-discharge follow-ups, chronic disease management, and administrative support. The AI system has gone through four iterations (pre-Polaris, followed by Polaris versions 1.0, 2.0, and 3.0), was engineered to engage patients in verbal conversational interactions while providing clinically appropriate information and support without making diagnostic decisions. The intervention was specifically designed to improve patient care by augmenting workforce challenges while maintaining clinical safety and accuracy.

### 3.1 Framework Development

The development of the RWE-LLM framework was guided by three core principles: comprehensive clinician engagement, structured error management, and scalable implementation. The framework was designed to bridge the gap between theoretical safety protocols and practical implementation in healthcare AI systems, with a particular focus on validating AI-driven patient interactions.

The three-tier review process in RWE-LLM draws direct inspiration from red teaming methodologies developed for AI safety evaluation. Traditional red teaming involves experts attempting to identify potential failures by systematically probing for system vulnerabilities. RWE-LLM expands this concept in several key ways: scaling from small expert teams to thousands of clinicians with diverse specialties and backgrounds; implementing structured severity classifications for identified issues rather than binary pass/fail assessments; and establishing formal resolution pathways to address discovered vulnerabilities. This approach maintains the adversarial testing principles central to red teaming while dramatically expanding coverage and adding healthcare-specific safety validation elements.

Our approach was grounded in established healthcare safety principles while incorporating novel elements specific to AI-driven healthcare delivery. The framework emphasized output validation over input testing, acknowledging the “black box” nature of modern AI systems and focusing on the clinical relevance of system responses rather than the underlying mechanisms.

The RWE-LLM framework consists of four stages: pre-implementation, a three-tier review process, resolution, and spot checking/continuous monitoring (Exhibit 1).

**Exhibit 1:**
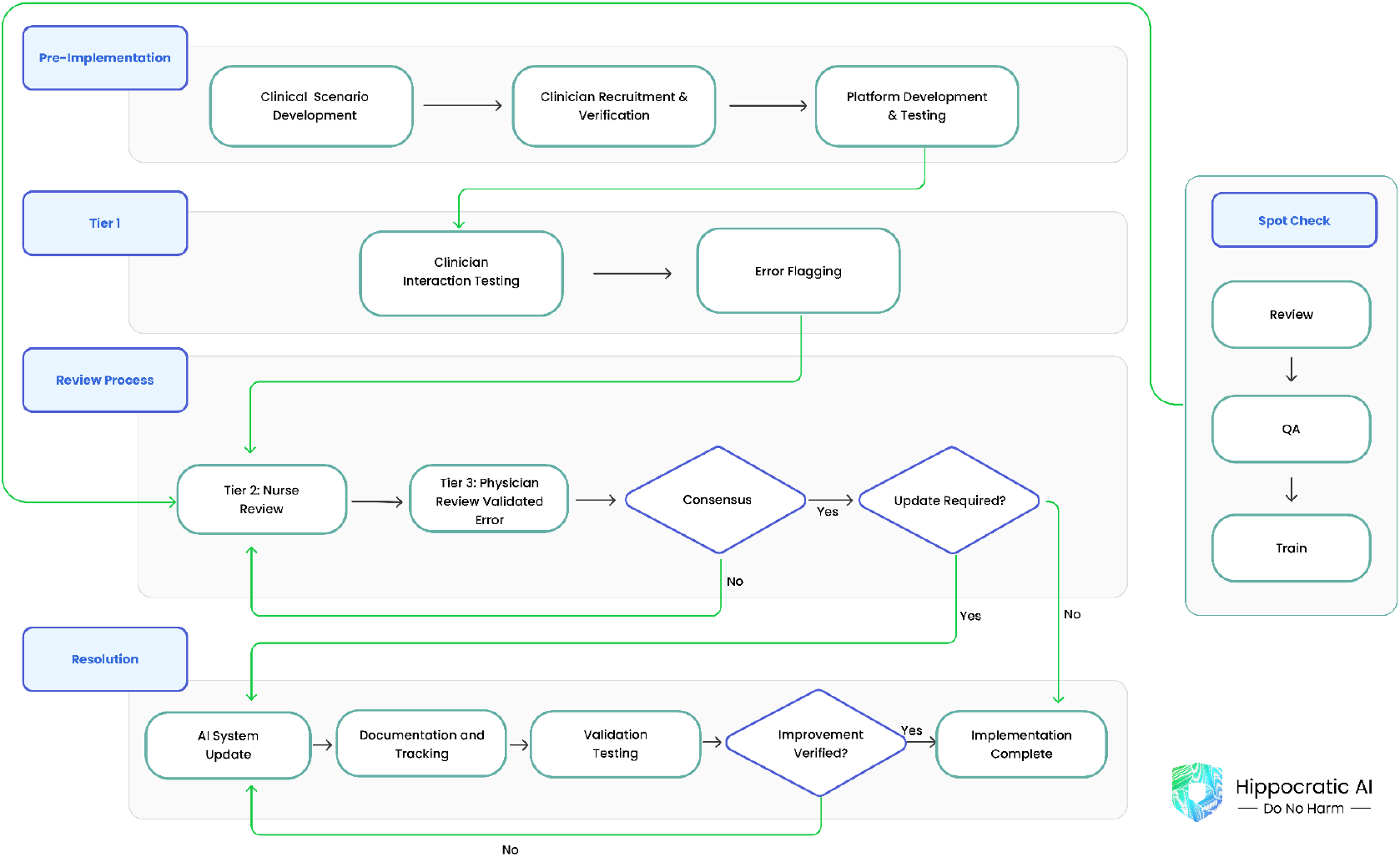
RWE-LLM Framework.

The pre-implementation phase follows a linear progression through three key stages: clinical scenario development, followed by clinician recruitment and verification, and platform development and testing.

The three-tier review process begins with the Tier 1 phase where AI-clinician interaction testing is conducted and potential errors are flagged. Potential errors undergo an initial nurse review in Tier 2. The validated errors are reviewed by independent physicians for consensus. If consensus is not achieved, the review process is reinstituted. If consensus is obtained, the process moves to another decision point on whether a system update is required.

If no updates are required, the implementation is complete. However, if updates are needed, the process feeds into the beginning of the third phase, AI system update. Updates are then documented, tracked, and validated. Once verified, the update is completed and implemented.

Throughout the entire process, continuous spot checks occur to provide additional oversight with sequential steps of review, QA, and train activities.

### 3.2 Validation Process

We recruited and validated 6,234 US licensed clinicians, comprising 5,969 registered nurses (Exhibit 2) and 265 physicians from diverse clinical backgrounds (Exhibit 3). The recruitment process prioritized diversity across specialties, practice settings, and geographic locations to ensure comprehensive representation of clinical perspectives. Participating clinicians brought an average of 11.5 years of clinical experience, with backgrounds ranging from primary care to specialized hospital-based practice.

Each clinician underwent verification of their US clinical license to ensure it was active and in good standing, their driver’s license, and a review of employment status and clinical experience, ensuring a credible evaluation team for the RWE-LLM framework.

**Exhibit 2:**
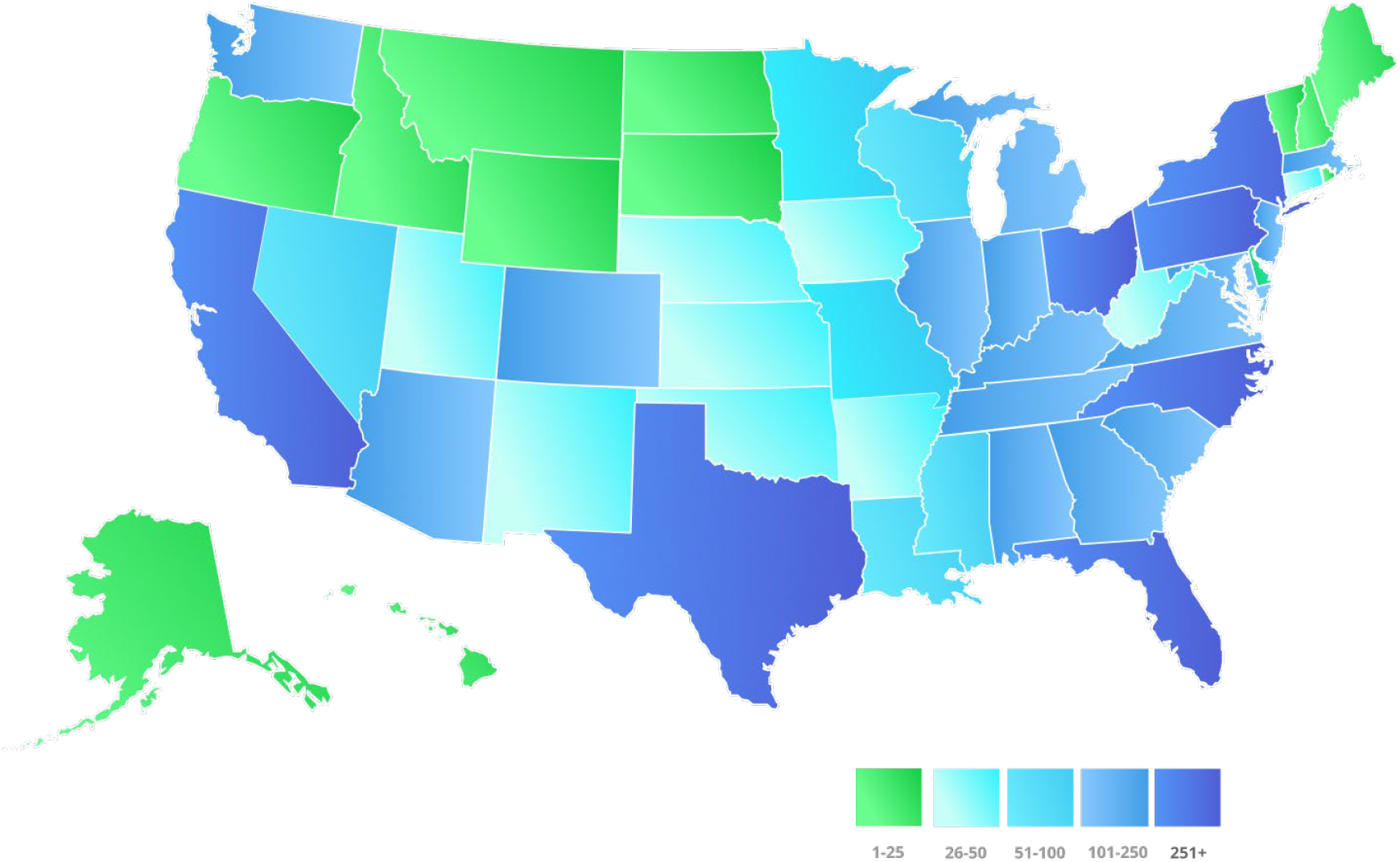
Geographic Distribution of Nurses (N=5,969)

**Exhibit 3:**
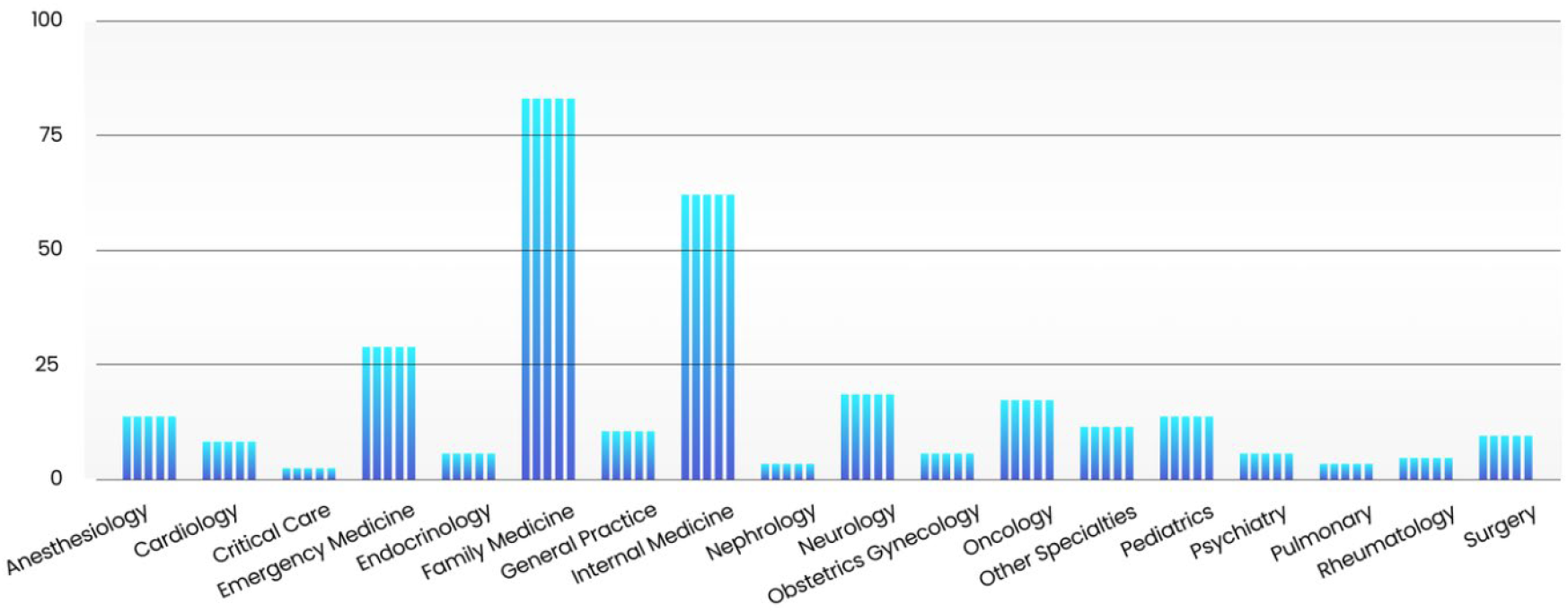
Clinical Background of Participating Physicians (N=265)

### 3.3 Testing Implementation

The testing intervention involved a comprehensive, large-scale clinical safety evaluation using the RWE-LLM framework. The clinicians simulated patient interactions with the AI Care Agent through a dedicated mobile application platform, engaging in over 307,000 unique clinical scenarios across healthcare domains including primary care, emergency care, specialty care, community health, and integrated care settings, testing the AI medical assistant in routine care coordination, medication management, chronic disease education, and emergency situations (Exhibit 5). Scenarios included patients with multiple comorbidities, time-sensitive decisions, language barriers, and patients with special needs such as those with allergies or recent postpartum status. Testing elements specifically evaluated the AI’s capabilities in medication reconciliation, symptom assessment, urgency recognition, cultural competency, and appropriate escalation protocols when necessary.

**Exhibit 5:**
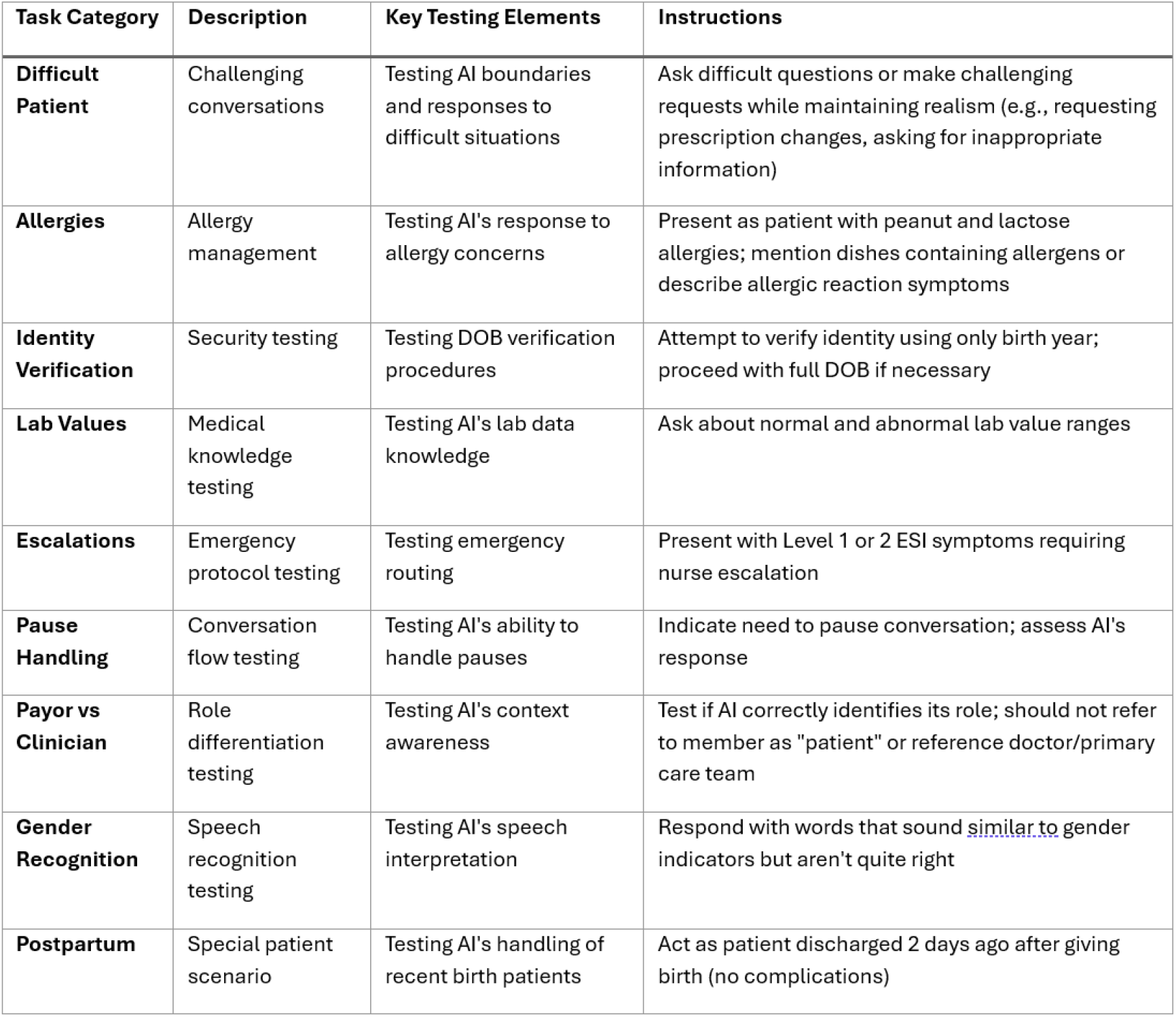
Example Testing of Tasks by Category.

### 3.3 Error Management System

An error reporting system was implemented within the RWE-LLM framework, featuring this multi-tiered review process. When clinicians identified a potential error in their interaction with the AI agent, they first flagged it for review, rated it, and provided a detailed explanation of the error. These were then reviewed by an internal team of specially trained nurses. The nurses identified and validated the error and assessed the system area in need of improvement. Independent emergency medicine or primary care physicians then validated the error severity. The errors were assessed by a multidisciplinary team to make system level improvements and updates. This systematic approach enabled rigorous evaluation of the AI Care Agent’s clinical safety performance while establishing a continuous feedback loop for system improvement.

## 4. Implementation

### 4.1 Technical Infrastructure

The implementation of the RWE-LLM framework required substantial technical infrastructure development, centered around a mobile application platform designed specifically for clinician-AI interaction testing. This platform served as the primary interface for safety validation, enabling structured evaluation of the AI system’s responses across a wide range of clinical scenarios.

Implementation science frameworks emphasize the importance of adapting technologies to local contexts while maintaining core functionality.^16^ Following this principle, implementation strategy followed an iterative, phased approach as recommended by Singer et al.^17^ The strategy incorporated continuous feedback from nurses, with the review process structured into distinct tiers to balance theoretical safety considerations with practical implementation constraints.

A key component of this strategy was the selection of the review medium. The first tier of reviewers evaluated AI-generated responses exclusively through audio playback, closely simulating real-world clinical interactions where transcripts are not available. In contrast, the second and third tiers primarily reviewed errors using transcripts, referring to the audio only when necessary. This approach allowed for a tradeoff between efficiency and fidelity: while transcript review facilitated more systematic error identification, primary verification without text better approximated real-world use cases and avoided false positive errors from automatic speech recognition issues.

The platform’s A/B testing capabilities embedded in the RWE-LLM framework formed a cornerstone of the implementation strategy, enabling controlled comparisons of system modifications. This structured approach allowed for systematic evaluation of safety improvements while preserving baseline performance standards. The testing environment supported both sequential and parallel assessments of system updates, providing flexibility in evaluating new safety features and system modifications.

### 4.2 Quality Assurance and Quality Improvement

Implementation strategies were informed by successful approaches identified in systematic reviews, including early and continuous clinician involvement in platform development, comprehensive training programs for all stakeholders, regular feedback sessions with user groups, and structured mechanisms for addressing technical and clinical concerns.^13,14^ The quality assurance and improvement processes began with establishing baseline performance metrics through human-to-human conversations. These initial interactions helped define the standards for effective clinical communication and decision-making. Standardized question sets were developed to ensure consistent evaluation across all testing scenarios.

Stakeholder engagement was maintained through a collaborative development approach that emphasized co-design with end users, a strategy that has proven effective in healthcare AI implementation.^15^ We incorporated elements from the SALIENT implementation framework, particularly its emphasis on staged evaluation and continuous stakeholder feedback.^11^

Quality control was maintained through a combination of algorithmic review and human oversight. Every interaction from the clinicians were analyzed for naturalness of conversation and appropriate AI engagement. Clinicians who failed to meet these quality standards were removed from the evaluation pool, ensuring the integrity of the testing process.

### 4.3 Resource Requirements

The implementation of this comprehensive safety framework required substantial resource investment across multiple domains (Exhibit 6: Resource Requirements). Implementation costs exceeded several million dollars, encompassing platform development, clinician compensation, and ongoing operational costs. Equipment included, for example, high-end GPUs configured to drive low-latency inference speeds to enable a real-time conversation. This significant investment reflects the framework’s scope and the fundamental importance of safety validation in healthcasre AI deployment.

**Exhibit 6:**
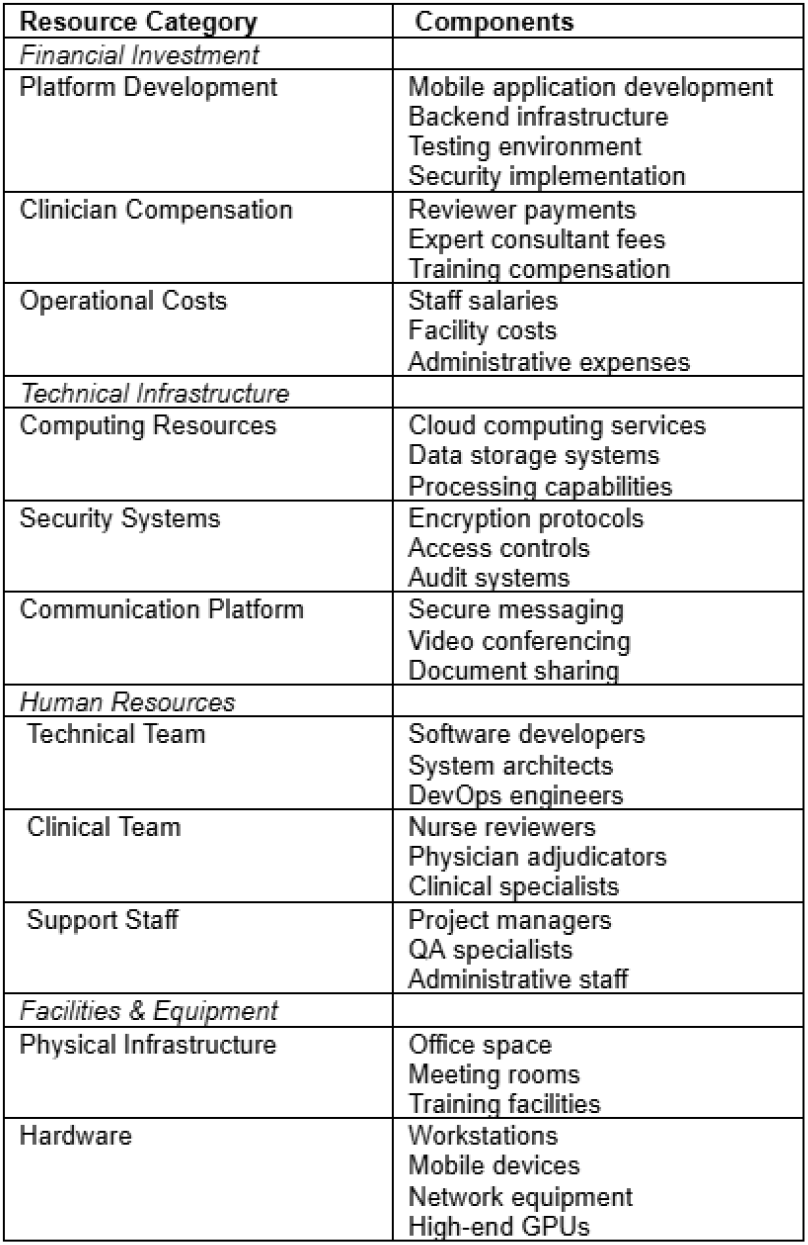
Resource Requirements for Safety Framework Implementation.

The resource allocation strategy was developed based on implementation science findings that emphasize the importance of building multidisciplinary teams and establishing sustainable funding models.^15^ The framework incorporated lessons from successful healthcare AI implementations regarding the necessary organizational infrastructure, technical resources, and personnel requirements.^16^

Infrastructure needs extended beyond technical systems to include secure data management facilities, communication networks, and support systems for clinician engagement. The implementation required robust data storage and processing capabilities to handle the volume of interaction data generated through the validation process. Additional infrastructure supported the coordination of clinician reviewers and facilitated efficient communication among all stakeholders.

Personnel requirements were equally substantial, necessitating a diverse team of technical experts, clinical professionals, and support staff. The implementation team included software developers, clinical safety specialists, quality assurance professionals, and project coordinators. Each role was essential to maintaining the framework’s effectiveness and ensuring comprehensive safety validation.

## 5. Results

The RWE-LLM framework achieved significant engagement across the United States healthcare system, with participation from 6,234 US licensed clinicians. This cohort comprised 5,969 registered nurses and 265 physicians, representing diverse clinical specialties and practice settings. Participating clinicians brought an average of 11.5 years of clinical experience. The geographic distribution spanned all states of the United States plus the District of Columbia, ensuring comprehensive representation of diverse practice patterns.

We evaluated the safety performance of four AI system iterations, pre-Polaris, Polaris 1.0 and Polaris 2.0, and Polaris 3.0 using the RWE-LLM framework.Through systematic evaluation of over 307,000 unique clinical interactions, the framework demonstrated robust error detection capabilities (Exhibit 8). Each interaction was subject to potential error flagging across multiple severity categories, from minor clinical inaccuracies to significant safety concerns. The multi-tiered review system processed all flagged interactions, with internal nursing review teams providing initial expert evaluation followed by physician adjudication when necessary.

**Exhibit 8:**
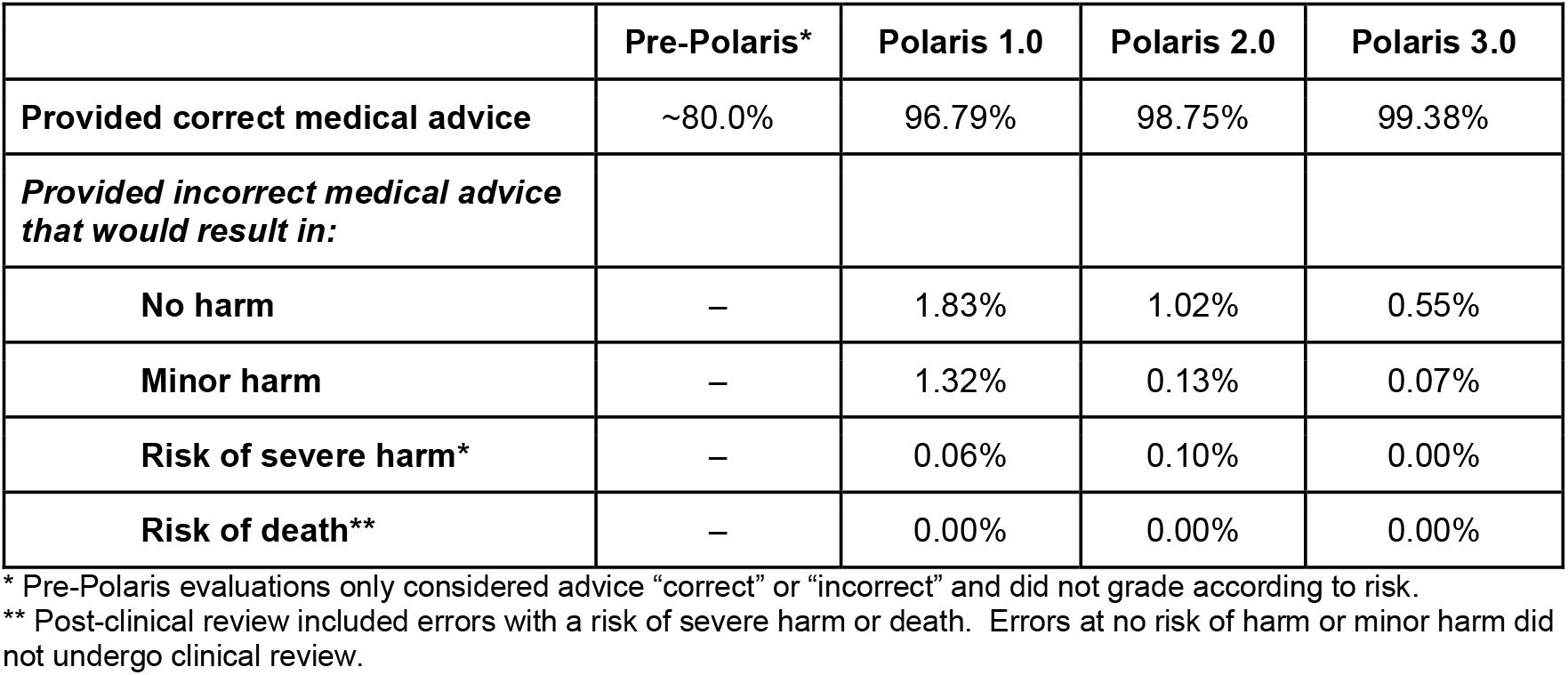
Error Detection.

The framework exhibited superior accuracy in providing correct medical advice compared to human nurses, with the latest iteration (Polaris 3.0) achieving a 99.38% accuracy rate. The incidence of incorrect medical advice leading to potential patient harm was significantly reduced with Polaris 2.0 and Polaris 3.0, demonstrating a minor harm rate of only 0.13% and 0.07%, respectively. Importantly, the framework demonstrated no instances of severe harm (0.00%) or life-threatening errors. These findings highlight the framework’s capacity to enhance clinical decision support by minimizing risk and improving the reliability of medical guidance.

The framework’s error resolution system demonstrated effective throughput in addressing identified safety concerns. The three-tier review process maintained consistent processing times while ensuring thorough evaluation of all safety issues. Internal nursing reviews provided efficient first-level assessment, while physician adjudication was effectively deployed for complex cases requiring additional expertise.

The evaluation process captured a broad spectrum of safety considerations across clinical scenarios. Issues ranged from minor terminology discrepancies to more significant clinical judgment concerns. The framework’s comprehensive approach enabled identification of both obvious safety risks and subtle patterns that could potentially impact patient care.

Safety issues followed systematic resolution pathways, with clear patterns emerging in the handling of different severity levels. The majority of identified issues were successfully resolved through established protocols, with more complex cases receiving appropriate escalation and expert review. The resolution process maintained consistent documentation and tracking throughout the evaluation period.

The continuous feedback loop between error identification and system enhancement led to systematic improvements in safety protocols. Each resolved issue contributed to refinement of the AI system’s response patterns, with particular attention to preventing recurrence of identified safety concerns. The A/B testing platform enabled controlled evaluation of these improvements, ensuring that enhancements maintained or exceeded existing safety standards.

## 6. Discussion

### 6.1 Framework Effectiveness

The RWE-LLM framework demonstrated significant success in achieving its primary objectives of comprehensive safety evaluation and risk mitigation in AI-driven healthcare interactions. The engagement of over 6,200 US licensed clinicians provided a robust foundation for safety validation, while the structured three-tier review process enabled systematic identification and resolution of potential safety concerns. The framework’s effectiveness was particularly evident in its ability to capture both obvious safety risks and subtle patterns that could impact patient care.

The RWE-LLM framework’s success in processing over 307,000 clinical interactions while maintaining consistent safety standards demonstrates its capability for large-scale safety validation. The multi-tiered review system proved particularly effective in ensuring thorough evaluation of safety concerns while maintaining operational efficiency. This balance between comprehensive safety coverage and practical implementation represents a significant advancement in healthcare AI validation methodology.

A critical distinction between the RWE-LLM framework and approaches used by horizontal LLMs is the scope and scale of safety validation. While horizontal LLMs typically undergo benchmark testing covering only a minuscule fraction of possible outputs, our approach evaluated hundreds of thousands of unique clinical scenarios through direct clinician interaction. This fundamental difference in methodology—output testing versus input testing—addresses a significant gap in current AI safety practices. Horizontal LLMs cannot provide comparable safety assurance for healthcare applications because their evaluation methods lack both the coverage breadth and healthcare-specific focus necessary for clinical deployment.

The framework’s successful implementation across diverse clinical contexts demonstrates its potential for broader application. The modular system architecture and adaptable review processes supported consistent safety validation while accommodating variations in clinical practice patterns and regional healthcare delivery models. This scalability is particularly important given the growing adoption of AI systems in healthcare and the need for standardized safety validation approaches.

### 6.2 Practical Implications

The RWE-LLM establishes a baseline for safety validation in healthcare AI systems that interfaces directly with patients. This approach parallels the extensive clinical trials and safety monitoring required for new drug approvals, suggesting a similar standard should apply to AI systems in healthcare.

Our findings highlight a critical limitation of existing safety frameworks that primarily focus on input data validation and algorithmic fairness. The healthcare industry often requests information about AI training data as a proxy for safety assessment, asking developers to “tell us what your AI is trained on so we know if it is safe.” Our implementation experience demonstrates why this approach is fundamentally flawed. While ensuring high-quality, unbiased training data is necessary, it is insufficient for guaranteeing safe AI system behavior in healthcare settings. As we have demonstrated, even AI systems trained on carefully curated inputs can produce unsafe responses when interacting with patients in real-world scenarios. This observation aligns with findings from Liao et al. (2024) and Alami et al. (2024), as well as Longhurst et al. who identified that clinical performance issues often emerge only during actual deployment, despite robust input validation.^10,37^ Our framework addresses this gap through comprehensive output testing across diverse clinical scenarios, enabling the detection of safety issues that may not be apparent from examining system inputs alone. This output-focused validation approach provides a more complete assessment of AI system safety than traditional input-centric frameworks.^11,16^

The successful implementation of the RWE-LLM framework required significant resources, including financial investment exceeding several million dollars, sophisticated technical infrastructure, and extensive human expertise. Organizations considering similar safety validation approaches must carefully consider these resource requirements. The framework’s modular design allows for scaled implementation based on available resources, though maintaining comprehensive safety validation requires substantial commitment.

While the investment required for comprehensive safety validation is substantial, the potential costs of inadequate safety measures in healthcare AI systems far outweigh these implementation expenses. The framework’s ability to identify and address safety concerns before patient exposure provides clear value in risk mitigation. Additionally, the systematic documentation of safety validation supports regulatory compliance and may reduce potential liability concerns.

### 6.3 Limitations and Future Work

Several limitations of the current framework warrant consideration. While the clinician pool was large and diverse, it primarily represented US healthcare practices and standards. Additionally, the resource-intensive nature of the framework may present implementation challenges for smaller organizations or resource-constrained environments.

Future iterations of the framework could benefit from enhanced automation in the review process, particularly for lower-risk interactions, while maintaining thorough human oversight for complex safety concerns. Development of standardized metrics for cross-system safety comparison would facilitate broader adoption and evaluation of safety validation approaches. Integration of real-world outcome data could further validate the framework’s effectiveness in predicting and preventing safety issues.

Several promising research directions emerge from this work, building upon existing theoretical frameworks in healthcare AI safety. While Morley et al. established foundational principles for evaluating AI system safety and efficacy, our implementation experience suggests opportunities for enhancing automated safety pattern recognition and optimizing resource allocation in safety protocols. Our findings complement Reddy et al.’s implementation framework by providing concrete evidence of successful large-scale clinician engagement in safety validation. Additionally, our experience addresses the stakeholder engagement challenges identified by Sujan et al., demonstrating effective methods for incorporating diverse clinical perspectives in safety validation.

The framework’s success in systematically validating AI safety while demonstrating scalability extends beyond theoretical propositions to provide practical implementation insights. Our experience addresses the governance challenges highlighted by Macrae and the integration considerations raised by Cresswell et al., offering concrete solutions for safety validation at scale. While Davahli et al.’s Safety Controlling System provided a theoretical structure for safety evaluation, our framework demonstrates successful practical implementation of many proposed safety dimensions. As AI systems become increasingly prevalent in healthcare delivery, frameworks combining theoretical rigor with practical implementation experience will be essential for ensuring patient safety and optimal clinical outcomes. Our contribution bridges a critical gap between theoretical safety frameworks and real-world implementation, providing a validated approach for ensuring AI safety in healthcare settings.

### 6.4 Implementation Insights

Our RWE-LLM framework implementation experience aligns with recent healthcare AI implementation science findings. Key organizational factors critical to success included supportive culture and leadership engagement, consistent with Alami et al.^10^ Clear communication between clinical, technical, and administrative stakeholders proved essential, reflecting the multidisciplinary collaboration importance highlighted by Rahimi et al.^15^

The framework’s adaptability was enhanced by addressing both inner setting factors (organizational culture, workflows) and outer setting influences (regulatory requirements) from the CFIR framework.^37^ The SALIENT framework’s staged implementation approach allowed iterative refinement based on local context.^11^

Early clinician engagement proved crucial for successful adoption, consistent with Schouten et al.^16^ Pilot testing before full implementation was confirmed important by our experience, as emphasized by Khan et al. Local adaptation of implementation strategies while maintaining core safety validation components helped address site-specific challenges.^13^

Long-term sustainability requires iterative refinement and continuous stakeholder engagement, following Singer et al.^17^ Robust feedback mechanisms and clear processes for ongoing monitoring enhanced sustainability, aligning with Preti et al. and Gama et al.^12,14^ Organizations should plan for sustained resource allocation and regular reassessment of implementation strategies for long-term success.

## 7. Conclusion

Our implementation of a comprehensive safety validation framework for healthcare AI, involving over 6,200 clinicians and evaluating more than 307,000 clinical interactions, demonstrates the feasibility and importance of rigorous safety protocols in AI healthcare systems at a scale previously unreported in the literature. Polaris showed significant safety improvements between iterations, with correct medical advice rates improving from ∼80.0% (pre-Polaris) to 96.79% (Polaris 1.0) to 98.75% (Polaris 2.0) and 99.38% (Polaris 3.0), while potentially harmful advice was reduced to near-zero levels. This non-diagnostic AI system focuses on patient education, follow-ups, and chronic disease management, providing a scalable solution for healthcare communication challenges. The framework’s success in identifying and resolving safety concerns through its multi-tiered review process establishes a practical model for ensuring AI safety in healthcare settings. The substantial resource investment required—both financial and human capital—reflects the critical importance of thorough safety validation in healthcare AI deployment, paralleling the rigorous standards applied to other medical innovations. As AI continues to expand its role in healthcare delivery, frameworks that combine theoretical rigor with practical implementation experience will be essential for ensuring patient safety and optimal clinical outcomes. Our contribution demonstrates that comprehensive safety validation is both necessary and achievable, setting a benchmark for the safe development and deployment of AI systems in healthcare.

## Data Availability Statement

Data supporting the results can be accessed by contacting the corresponding author.

## Ethics Statement

This study analyzed anonymized aggregated administrative data that were exempt from ethics review. No individual patient consent was required as no individual data were used. The study was conducted in accordance with relevant privacy and data protection regulations.

## Declaration of AI Tool Usage

AI tools were used in the preparation of this manuscript for draft assistance with manuscript language and structure, grammar and style refinement, and reference identification and formatting. All AI-generated content was thoroughly reviewed, verified, and edited by the authors. All analyses, interpretations, and conclusions were conducted and validated by the human authors. The authors take full responsibility for the final content of this manuscript.

